# Adherence of SARS-CoV-2 seroepidemiologic studies to the ROSES-S reporting guideline during the COVID-19 pandemic

**DOI:** 10.1101/2023.06.02.23290895

**Authors:** Brianna Cheng, Emma Loeschnik, Anabel Selemon, Reza Hosseini, Jane Yuan, Harriet Ware, Xiaomeng Ma, Christian Cao, Isabel Bergeri, Lorenzo Subissi, Hannah C. Lewis, Tyler Williamson, Paul Ronksley, Rahul K. Arora, Mairead Whelan, Niklas Bobrovitz

## Abstract

**Background:** Complete reporting of seroepidemiologic studies (e.g. sampling and measurement methods, immunoassay characteristics) are critical to their interpretation, comparison, and utility in evidence synthesis. The Reporting of Seroepidemiologic studies—SARS_JCoV_J2 (ROSES-S) guideline is a reporting checklist that aims to improve the quality and transparency of reporting in SARS-CoV-2 seroepidemiological studies. While the synthesis of seroepidemiologic studies played a crucial role in public health decision-making during the COVID-19 pandemic, adherence of SARS-CoV-2 seroepidemiologic studies to the ROSES-S guideline has not yet been evaluated.

**Objectives:** To evaluate the completeness of SARS-CoV-2 seroepidemiologic study reporting over the first two years of the COVID-19 pandemic by assessing adherence to the ROSES-S reporting guideline, determine whether publication of the ROSES-S guideline was associated with changes in reporting completeness, and identify study characteristics associated with reporting completeness.

**Methods:** A stratified random sample of SARS-CoV-2 seroepidemiologic studies from the SeroTracker living systematic review database was evaluated for adherence to the ROSES-S guideline. We categorized study adherence to each reporting item in the guideline as “reported”, “not reported”, or “not applicable”. For each reporting item we calculated the percentage of studies that were adherent. We also calculated the median and interquartile range (IQR) adherence across all items and by item domain. Piecewise and multivariable beta regression analyses were used to determine whether publication date of the ROSES-S guideline was associated with changes in the overall adherence scores and to identify study characteristics associated with overall adherence scores.

**Results:** 199 studies were included and analyzed. The median adherence to reporting items was 48.1% (IQR 40.0%–55.2%) per study. Adherence to reporting items ranged from 8.8% to 72.7% per study. The laboratory methods domain (e.g. description of testing algorithm) had the lowest median adherence (33.3% [IQR 25.0%–41.7%%]), while the discussion domain had the highest median adherence (75.0% [IQR 50.0%–100.0%])). There were no significant changes in reporting adherence to ROSES-S before and after guideline publication. Article publication source (p<0.001), study risk of bias (p=0.001), and sampling method (p=0.004) were significantly associated with adherence to the ROSES-S guideline.

**Conclusions:** The completeness of reporting in SARS-CoV-2 seroepidemiologic studies was suboptimal, especially in laboratory methods, and was associated with key study characteristics. Publication of the ROSES-S guideline was not associated with changes in reporting practices. Given that reporting is necessary to improve the standardization and utility of seroprevalence data in evidence synthesis, authors should improve adherence to the ROSES-S guideline with support from stakeholders.

## Introduction

Measuring the prevalence of antibodies against SARS-CoV-2 in population blood and serum samples (seroprevalence) provides valuable estimates of true population infection, including infections in asymptomatic individuals or those who aren’t able to acquire diagnostic services.^1^ This makes serosurveillance a valuable tool for pandemic decision-making. Seroepidemiologic studies must be well reported to be useful, however, as information about how seroprevalence was measured (e.g. immunoassay characteristics and validation), sampling methods, and statistical techniques affects study interpretation, comparison, and application.^2^

Inconsistent reporting of study methods and results was evident during the first year of the COVID-19 pandemic,^3^ despite the existence of guidelines for study designs typically used to estimate seroprevalence (e.g. the “Strengthening the Reporting of Observational Studies in Epidemiology” (STROBE) guidelines).^4^ Reporting guidelines were also available for specific domains of epidemiological research, such as observational infectious disease studies (e.g. the “Strengthening the Reporting of Molecular Epidemiology for Infectious Diseases” [STROME-ID] statement) and influenza seroepidemiology (e.g. the CONSISE “Reporting Of Sero-Epidemiologic Studies for Influenza” [ROSES-I]) statement. Previous studies suggest that use of these types of guidelines improves reporting,^5, 6^ yet adherence to such guidelines appears suboptimal. For instance, estimates of adherence to STROBE’s 22 reporting criteria have ranged from 51.4% to 76.5%,^7–10^ with similar estimates of adherence (50%) to STROME-ID.^11^

In June 2021, the World Health Organization (WHO) published the “Reporting of Sero-Epidemiologic Studies - SARS-CoV-2” (ROSES-S) guideline as a content-specific extension of the STROBE checklist. ROSES-S is a checklist of 22 criteria for reporting SARS-CoV-2 seroepidemiologic studies regardless of study design.^12^

To better understand reporting completeness among SARS-CoV-2 seroepidemiologic studies and determine whether ROSES-S has changed practice, we conducted a sub-study within a living systematic review with the following aims: i) quantify the completeness of reporting among SARS-CoV-2 seroepidemiologic studies by assessing adherence to the ROSES-S reporting guideline; (ii) assess whether specific study characteristics were associated with adherence to the ROSES-S guideline; (iii) determine whether implementation of the ROSES-S guideline was associated with changes in the reporting completeness of SARS-CoV-2 seroepidemiologic studies.

## Methods

### Search Strategy and Eligibility Criteria

This study was embedded within SeroTracker’s living global systematic review and meta-analysis of SARS-CoV-2 seroepidemiologic studies (PROSPERO [CRD42020183634])^13^, the results of which are available on a public dashboard and data platform.^14^ An in-depth description of the methods, including a full search strategy, inclusion and exclusion criteria, and screening process, has been previously published.^14, 15^ Briefly, a search of electronic databases, grey literature, and news media captured published and pre-print studies, as well as institutional reports from January 1, 2020 to December 31, 2022. Direct submissions of data were also invited at SeroTracker.com.

A seroepidemiologic study was defined as estimating the prevalence of SARS-CoV-2 antibodies in humans through the collection and testing of serum (or proxy such as oral fluid) specimens from a defined sample frame over a specified period of time. Eligible study designs included cross sectional studies, repeated cross-sectional studies, and cohort studies. Studies of all languages were included if they reported the number of participants, sampling end date and week, geographic location of sampling, and a prevalence estimate. Studies in languages other than English, French, Spanish, Romanian, Persian, and Chinese were translated via Google translate machine translation.

Studies were screened using Covidence systematic review manager^16^ via a two-stage screening process by two independent reviewers. The first stage determined inclusion based on review of study title and/or abstract. The second stage determined inclusion by review of the full text. A third team member resolved discrepancies by arbitration.

### Study Selection

We included a random time-stratified sample (n=199) of peer-reviewed, pre-print studies, and grey literature (government and institutional reports), from March 2020 to December 2022 to capture studies before and after the publication of the ROSES-S guideline (June 2021). The sample size was determined based on a precision level of ± 7% with 95% confidence, and a 60% estimated adherence to checklist items.^11, 17^ To ensure a comparable sample size before and after the ROSES-S guideline’s publication date, we created strata of six month discretized time periods and randomly sampled 34 studies from each strata. If a given month had only one included study then we conducted additional random sampling of studies from that month to select an additional study - bringing the total to two.

### Data Extraction

The following data was extracted by one independent reviewer using an Airtable spreadsheet tool (2022) and verified by a second reviewer from the SeroTracker team: article publication source, study design, publication month, article endorsement of ROSES-S or any other reporting guidelines, sampling frame, sampling method, sample size, geographic scope (local, regional, national), WHO Humanitarian Response Plan (HRP) status^18^, WHO Unity protocol alignment^19^, SARS-CoV-2 antibody seroprevalence estimate(s), and whether the article cited the ROSES-S guideline or other reporting guidelines. Risk of bias (low, moderate, high) was evaluated by two independent reviewers using a modified version of the Joanna Briggs institute checklist.^20, 21^ Further details on extraction methods can be found in a previous publication.^22^

### Reporting Guideline Adherence Scoring System

The ROSES-S guideline scoring sheet split each checklist criteria into its component paragraphs, with each paragraph containing specific and related reporting information (Supplementary file 1). As a result, a total of 68 items were scored independently. Only the second component of criteria 5 (“Describe the timing of the biological sampling in relation to the disease epidemiology in the study population [the beginning, peak, and end of virus transmission], Describe any vaccination efforts that have been undertaken”) was further split into two items (items 5.2 and 5.3), as vaccination was not relevant in the beginning of the pandemic.

To ensure inter-rater agreement, raters (AS, BC, EL) piloted ten eligible studies; discordances were discussed with arbitration by a co-creator of the ROSES-S guideline (NB). Scoring was completed by categorizing each statement as “reported” or “not reported”. A “not applicable” (N/A) option was possible for 22/68 (32.4%) items related to specific study designs or that were not relevant at all stages of the pandemic. For example, items pertaining to case-control studies were automatically “not applicable” given that the case-control study design was not eligible for inclusion in our study. N/A scores were not assigned a numeric value and, therefore, were not included in the denominator when calculating adherence proportions. Where there were multiple criterion embedded in a single reporting item that was not further broken down in the scoring sheet, the item was scored as “reported” if any of the criteria were satisfied. For each study, the adherence scores for each of the six domains were calculated, as well as an overall adherence score across all items.

### Statistical Analysis

Study characteristics were summarized using descriptive statistics. We calculated the percentage of studies adhering to each item of the ROSES-S scoring sheet, as well as the median percentage and interquartile range (IQR) of adherence across all items and for each domain (i.e. title, abstract, and introduction; epidemiological methods; laboratory methods; results; discussion; other information) and for each study characteristic (article publication source, study design, publication month, sampling frame, sampling method, sample size, overall risk of bias, geographic scope, WHO HRP status, WHO Unity protocol alignment, and whether the study cited ROSES-S or other reporting guidelines). Overall percentage of adherence and percentage adherence by domain were visualized with violin plots. A publication time lag of 154 days was used; this was the estimated median time between the last date of participant sampling and the publication date of SARS-CoV-2 seroepidemiologic studies during that time period, irrespective of publication type.^23^ Three time periods were used; pre-ROSES-S publication (March 1, 2020– June 26, 2021), 0-154 days post-ROSES-S publication (June 27, 2021–November 27, 2021), and 155-553 days post-ROSES-S publication (November 28, 2021-December 31, 2022). Risk of bias evaluations were summated using percentages.

Beta regression was used to investigate the association of study characteristics and publication of the ROSES-S guideline with total adherence scores. A univariable continuous piecewise model with two knots, one at the date of ROSES-S publication and the other 154 days later, was constructed to assess the total adherence scores over time and was plotted to conduct a visual inspection of trends. A multivariable model was also built to explore all candidate study characteristic predictors. Marginal effects were calculated for the final model to facilitate the interpretation of model coefficients. All adherence scores and predicted score changes from the beta regression models are reported as percentages. See Supplementary file 2 for more details. Analysis was conducted using the betareg (version [v] 3.1.4)^24^, lmtest (v 0.9.40)^25^, MuMIn (v 1.47.5)^26^, and mfx (v 1.2.2)^27^ packages in R (v 4.2.2).

## Results

199 articles were included and analyzed (Appendix Figure 1) (Supplementary file 3). Study characteristics are reported in Table 1. Included studies were mostly published in peer-reviewed journals (68.3%), used one-time cross-sectional survey design (74.9%) and were sampled from the general population (52.8%). The studies mostly used a non-probability sampling method (71.3%) and incorporated a sample size of 1,000 participants or greater (46.2%). The majority of studies were local in geographic scope (61.3%) and originated from geographic locations without HRP status (81.4%). Most studies were not aligned to the WHO Unity Studies protocol (77.9%) and did not cite any reporting guideline (97%), with none citing ROSES-S. Lastly, articles were assessed to have a high risk of bias (61.8%), with risk of bias item 8b (“Was there appropriate adjustment for population characteristics?”) the least fulfilled (12.6%) (Appendix Table 1).

**Figure 1:**
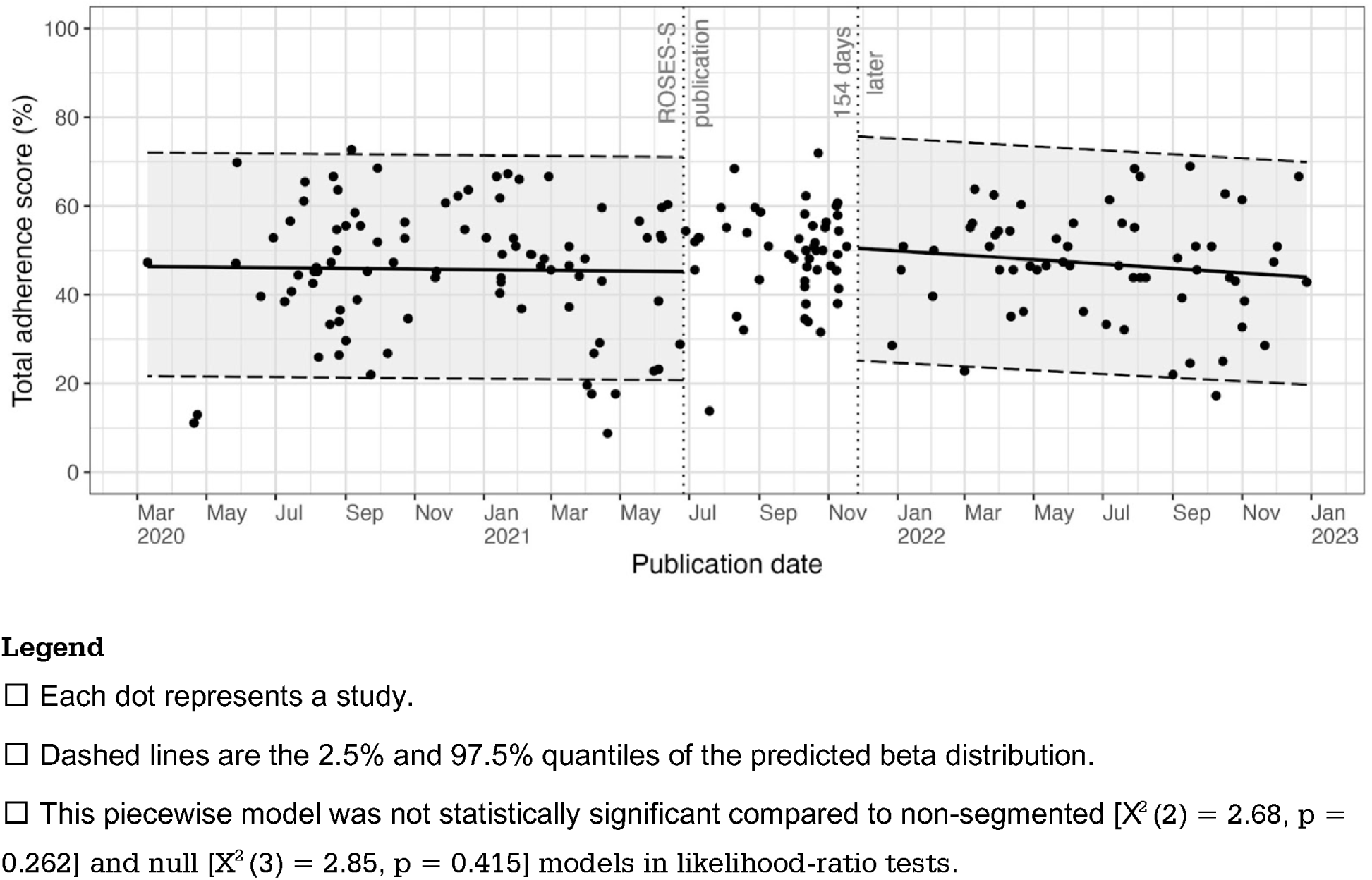
Adherence to ROSES-S over time

**Table 1:** Summary characteristics of included articles (n=199)

| **Variable** | **Studies, n (%)** |
| --- | --- |
| **N** | 199 |
| **Source Type, n/N (%)** |  |
| Journal article (Peer-Reviewed) | 136/199 (68.3) |
| Pre-print | 39/199 (19.6) |
| Institutional report | 17/199 (8.5) |
| Conference, Abstract or News | 7/199 (3.5) |
| **Study Design, n/N (%)** |  |
| Cross-sectional survey | 149/199 (74.9) |
| Repeated cross-sectional survey | 21/199 (10.6) |
| Cohort study | 29/199 (14.6) |
| **Sampling Frame<sup>†</sup>, n/N (%)** |  |
| General population | 105/199 (52.8) |
| Special population | 94/199 (47.2) |
| **Sampling Method, n/N (%)** |  |
| Non-Probability | 142/199 (71.3) |
| Probability | 40/199 (20.1) |
| Unclear | 17/199 (8.5) |
| **Sample Size, n/N (%)** |  |
| <500 | 63/199 (31.7) |
| 500-999 | 44/199 (22.1) |
| 1000+ | 92/199 (46.2) |
| **Overall JBI, n/N (%)** |  |
| High | 123/199 (61.8) |
| Moderate | 66/199 (33.2) |
| Low | 10/199 (5.0) |
| **Geographic Scope, n/N (%)** |  |
| National | 42/199 (21.1) |
| Regional | 35/199 (17.6) |
| Local | 122/199 (61.3) |
| **HRP Status, n/N (%)** |  |
| HRP | 37/199 (18.6) |
| Non-HRP | 162/199 (81.4) |
| **WHO Unity Protocol<sup>†</sup>, n/N (%)** |  |
| Unity-Aligned | 44/199 (22.1) |
| Not Unity-Aligned | 155/199 (77.9) |
| **Citation of Reporting Guideline, n/N (%)** |  |
| STROBE | 5/199 (2.5) |
| TREND | 1/199 (0.5) |
| ROSES-S | 0/199 (0) |

<sup>†</sup> General population includes the following sampling frames: household and community samples, blood donors, residual sera, persons living in slums, pregnant or parturient women, and representative patient populations.
<sup>‡</sup> WHO Unity aligned: studies aligned with the WHO Unity protocol.
Abbreviations: HRP, Humanitarian Response Plan; STROBE, Strengthening the Reporting of Observational Studies in Epidemiology; TREND, Transparent Reporting of Evaluations with Nonrandomized Designs.

Adherence of study reporting to ROSES-S items are reported in Table 2. The median adherence to ROSES-S reporting items was 48.1% (IQR 40.0%–55.2%) per study. Adherence ranged from 8.8% to 72.7% per study. Overall, the most frequently reported ROSES-S items were “State the interval between sequential biological samples (serial cross-sectional or longitudinal studies), or specify whether only a single sample was collected (cross-sectional study)” (item 5.5: 197/199 [99.0%]), and “For a crosslJsectional study, report the numbers of outcome events or summary measures” (item 15.3: 160/163 [98.2%]). The least frequently reported ROSES-S items were “For a cohort study, explain how loss to followlJup was addressed, if applicable” (item 12.5: 0/14 [0%]) and “Specify laboratory biosafety conditions” (item 13.10): 2/198 [1.0%]).

**Table 2:**
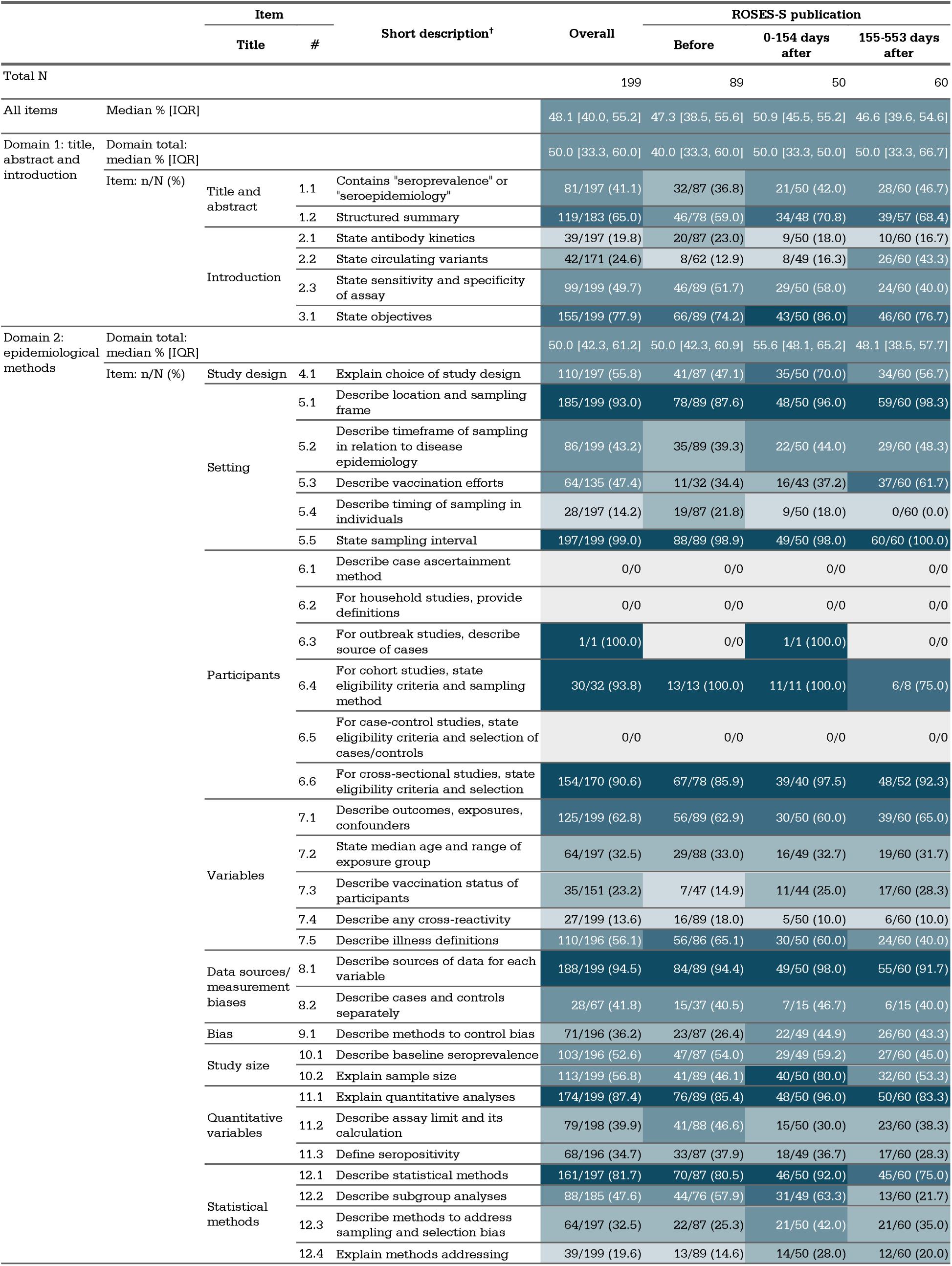
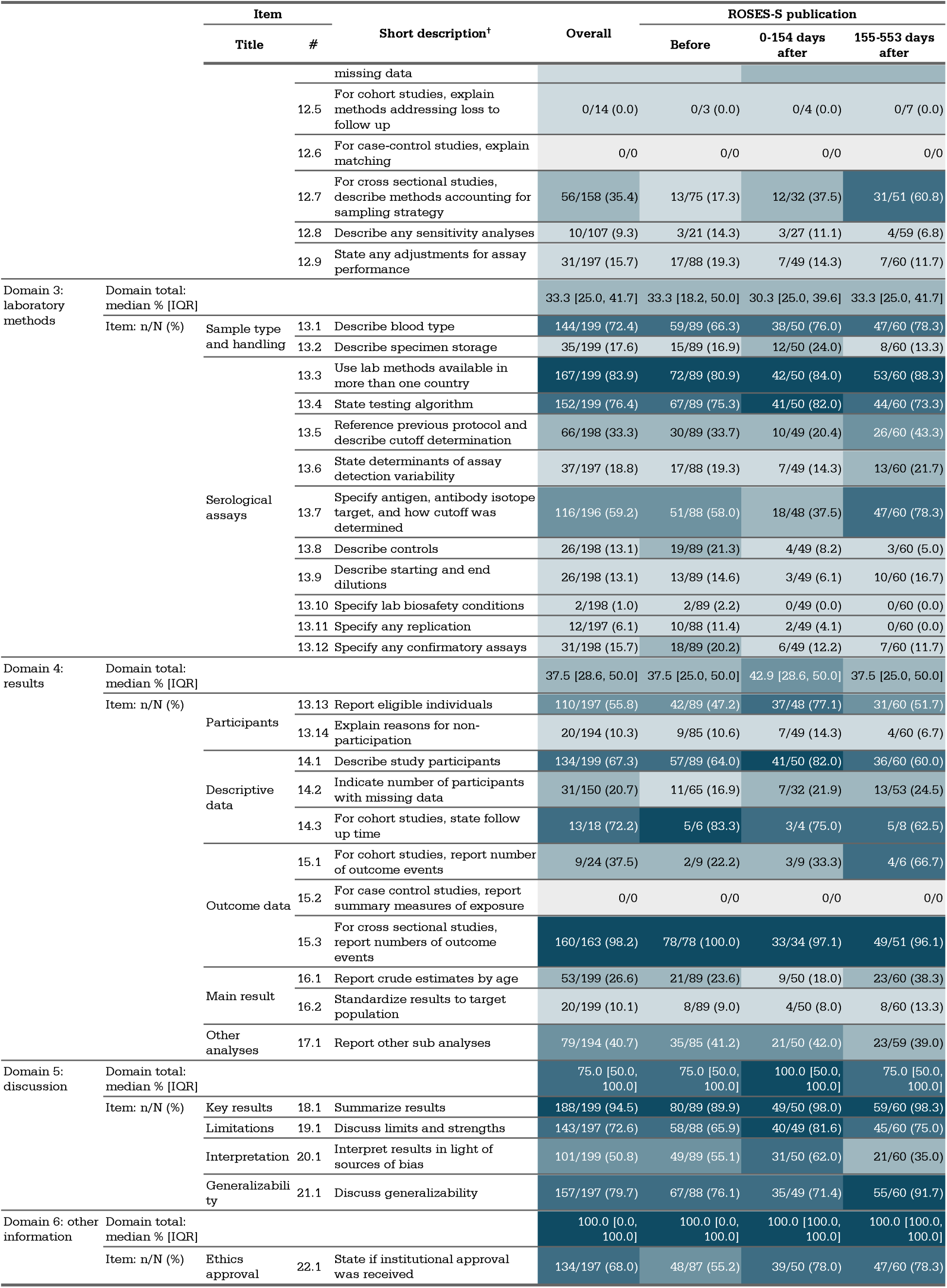
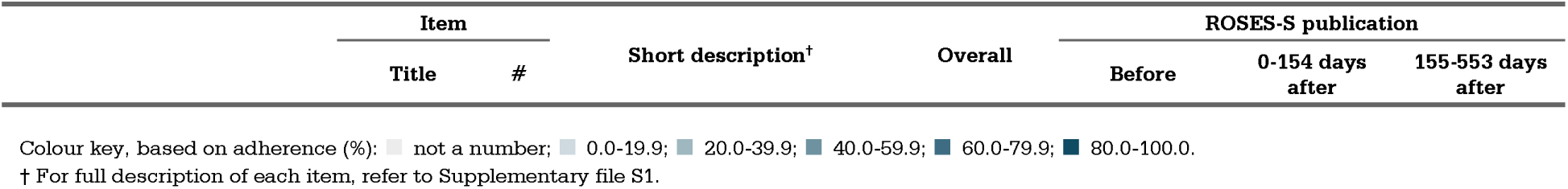
Adherence to ROSES-S overall and pre/post ROSES-S publication

Adherence scores for the ROSES-S domains varied (Table 2 and Appendix Figure 2). “Laboratory methods” (domain 3) was the domain with the least frequently reported item(s) with a median of 33.3% (IQR 25.0%–41.7%), followed by “Results” (domain 4) (37.5% [IQR 28.6%– 50.0%]) (Table 2). Discussion was considered the domain with the most frequently reported item(s), with a median of 75.0% (IQR 50.0%–100.0%). “Other information” (domain 6) was reported in 100% 100% of studies but only consisted of one item (item 22.1).

Reporting varied by study characteristics (Appendix Table 2). The highest median adherence to the ROSES-S items was among pre-print publications (52.6% [IQR 45.9%– 61.5%]), and studies that used cohort study design (50.9% [IQR 43.9%-56.1%). Studies that sampled from special populations (49.1% [IQR 43.9%–56.3%]), used a probability sampling method (51.3% [IQR 45.6%–59.6%]), incorporated a sample size of 1,000+ participants (50% [IQR 43.6%–56.4%]), were assessed to have a moderate risk of bias (52.7% [IQR 45.3%– 59.4%]), were regional in geographic scope (51.9% [IQR 43.9%–61.3%]), and originated from geographic locations with HRP status (51.9% [IQR 42.9%–56.1%]) had the highest median adherence to the ROSES-S items. Studies that were aligned to the WHO Unity Studies protocol (49.1% [IQR 42.8%–53.7%]), and cited any reporting guideline (STROBE and TREND) (57% [IQR 48.9%–59.2%]) also had the highest median adherence to the ROSES-S items.

Median adherence to the ROSES-S items was similar before and after the publication of the ROSES-S guideline (Table 2). Median adherence was 47.3% (IQR 38.5%–55.6%) prior to ROSES-S publication, 50.9% (IQR 45.5%–55.2%) 0-154 days after publication, and 46.6% (IQR 39.6%–54.6%) 155-553 days after publication. Adherence to items fluctuated over time, increasing for some items after the publication of the ROSES-S guideline and decreasing for others. For example, reporting of “For a cross-sectional study, describe analytical methods taking account of sampling strategy, if applicable” (item 12.7) increased from 17.3% (13/75 studies) pre-ROSES-S to 60.8% (31/51 studies) 155-553 days post-ROSES-S. In contrast, reporting of “Describe illness definitions and methods for ascertaining the presence or absence of clinical illness in subjects” (item 7.5) decreased from 65.1% (56/86 studies) pre-ROSES-S to 40% (24/60 studies) 155-553 days post-ROSES-S. Piecewise time series analysis showed no significant changes in reporting after the publication of the ROSES-S guideline (Figure 1).

Multivariable analysis for beta regression is presented in Table 3. Overall reporting adherence was significantly associated with article publication source, study risk of bias (RoB), and sampling method. Institutional reports and conference abstracts or news articles had considerably lower mean total adherence to ROSES-S, compared to published peer-reviewed journal articles, respectively at -14.0% (−19.4 – -8.6) and -29% (−35.1 – -22.9). Mean total adherence to ROSES-S was 5.8% (2.3% – 9.3%) and 7.8% (0.3% – 15.2%) higher for studies with moderate and low RoB, respectively, compared to those with a high RoB. Furthermore, studies that used an unclear sampling method had a -9.4% (−15.8% – -2.9%) lower mean total adherence to ROSES-S compared to those that used a non-probability sampling method. Univariable analyses also showed special population studies had higher adherence scores compared to general population studies 4.2% (0.5% – 7.8) (Appendix Table 3).

**Table 3:** Multivariable beta regression identifying factors associated with reporting completeness

| Component | Variable |  | Model coefficients |  |  | Marginal effects (%) |  |  |
| --- | --- | --- | --- | --- | --- | --- | --- | --- |
|  |  |  | Estimate (95% CI) | p |  | Estimate (95 % CI) | p |  |
| Mean | (Intercept) | (Intercept) | -0.12 (-0.21 - -0.04) | 0.006 | ** |  |  |  |
|  | Source type | Peer-reviewed journal article | Reference |  |  | Reference |  |  |
|  |  | Preprint | 0.14 ( 0.00 - 0.28) | 0.051 | . | 3.54 ( -0.04 - 7.12) | 0.051 | . |
|  |  | Institutional report | -0.58 (-0.82 - -0.34) | <0.001 | *** | -13.98 (-19.41 - -8.55) | <0.001 | *** |
|  |  | Conference abstract or news | -1.37 (-1.76 - -0.99) | <0.001 | *** | -29.03 (-35.13 - -22.94) | <0.001 | *** |
|  | Overall risk of bias | High | Reference |  |  | Reference |  |  |
|  |  | Moderate | 0.23 ( 0.09 - 0.37) | 0.001 | ** | 5.81 ( 2.32 - 9.31) | 0.001 | ** |
|  |  | Low | 0.31 ( 0.01 - 0.61) | 0.041 | * | 7.77 ( 0.32 - 15.22) | 0.040 | * |
|  | Sampling method | Non-probability | Reference |  |  | Reference |  |  |
|  |  | Probability | 0.00 (-0.15 - 0.16) | 0.955 |  | 0.11 ( -3.86 - 4.08) | 0.955 |  |
|  |  | Unclear | -0.38 (-0.66 - -0.11) | 0.006 | ** | -9.37 (-15.84 - -2.90) | 0.004 | ** |
| Precision | (Intercept) | (Intercept) | 3.05 (-0.21 - -0.04) | <0.001 | *** |  |  |  |
|  | Sampling method | Non-probability | Reference |  |  |  |  |  |
|  |  | Probability | 0.56 (-0.15 - 0.16) | 0.025 | * |  |  |  |
|  |  | Unclear | -0.38 (-0.66 - -0.11) | 0.277 |  |  |  |  |

## Discussion

This systematic review sub-study examined the completeness and associated factors of reporting in SARS-CoV-2 seroepidemiologic studies published in the first two years of the COVID-19 pandemic as determined by adherence to the ROSES-S guideline. Reporting completeness was found to be suboptimal. The median adherence to ROSES-S items was 48.1% per study, which is comparable to estimates from studies evaluating adherence to STROBE and STROME-ID.^7–11^

There was variation in the completeness of reporting for different reporting items. Domains of the ROSES-S guideline with the least frequently reported items were laboratory methods and results, while the least reported individual items related to serological assays and statistical methods. This aligns with evaluations of STROBE, where key methodological items seem to be consistently underreported (e.g. sample size estimation, addressing missing data, addressing loss to follow-up).^28^ This trend persists even in evidence synthesis citing more heavily journal endorsed and established guidelines, such as the “Preferred Reporting Items for Systematic reviews and Meta-Analyses” (PRISMA) statement^29^, with one review finding fewer than 67% of systematic reviews adhering to items relating to protocol and registration, search, data items, risk of bias across and within studies, and additional analyses.^30^

Publication of the ROSES-S guideline was not associated with a change in reporting completeness, suggesting that the guideline has not had an impact on reporting practices to date. Other studies using segmented regression to evaluate the impact of the STROBE guideline publication on reporting have similarly found no significant association.^8^ This contrasts with evaluations of the “Consolidated Standards of Reporting Trials” (CONSORT) guideline,^31^ which was originally published in 1996 and last updated in 2001,^32^ as well as the PRISMA statement. According to PRISMA, from publication in 2010 to 2016, item reporting adherence increased in systematic reviews, compared to systematic reviews published from 1989 to 2016.^30^ This suggests that a longer follow-up time may be needed for awareness and uptake of the ROSES-S guideline, which was published in 2021. This is reinforced by our finding that only 3.7% of peer reviewed studies cited a reporting guideline, none of which were ROSES-S, suggesting a lack of awareness of the ROSES-S guideline.

Complete reporting of methodological items is necessary to inform appropriate adjustments of estimates based on differences in study methodology. Methodological heterogeneity has been observed in previous syntheses of SARS-CoV-2 seroepidemiologic studies and has been associated with notable differences in estimates.^30, 31^ For example, in one study, failing to account for methodological differences between serosurveys led to variation in estimates as high as 16%.^31^ In instances where methods are not clearly reported, adjustment may not be possible, and this may affect the accuracy, comparability, and generalizability of synthesized seroprevalence estimates.

Complete reporting of items related to sampling and measurement methods are particularly important given their impact on study risk of bias (RoB).^21^ ROSES-S items, such as those related to sample recruitment, size, and retention, as well as immunoassay validation and testing algorithm, inform judgements of selection bias and measurement bias, respectively.^21^ For instance, incomplete reporting of immunoassay validation details may contribute to misleading results as it limits the opportunity for adjustment of seroprevalence estimates for poor test performance, which has been shown to heavily influence seroprevalence estimates.^31^ Incomplete reporting thus directly affects RoB assessment and creates the potential for misleading meta-analyzed estimates, hindering the utility of individual studies and evidence synthesis.

Interventions to improve adherence to reporting guidelines have been proposed and tested. For instance, there could be greater encouragement of use of the guidelines, such as integrating checklist writing tools in the submission process (e.g. COBWEB), which have improved overall reporting completeness for randomized controlled trials.^33^ Although journal endorsement of reporting guidelines seems to impact adherence,^6, 32, 34^ it can be ineffective without enforcement. Verifying adherence to a guideline throughout the peer-review process could also be considered. Editors may ask authors to flag non-adherence to a given guideline. When such an intervention was evaluated using the CONSORT guideline, improvements in reporting were demonstrated.^35^ Peer-review tools, such as StatReviewer, that automatically check guideline adherence against manuscripts may also improve adherence.^36^

There are other potential solutions specifically tailored for the reporting of serosurveillance data. For instance, reporting guidelines could be included in epidemiological study protocols, like the UNITY studies protocol for COVID-19^19^, to increase awareness, and to place emphasis on methodological quality and reporting quality at the outset of conducting a study. Reporting completeness and guidelines should also be considered by stakeholders establishing and maintaining serological surveillance networks in preparation for the next pandemic.

We found that institutional reports, news and media reports, and conferences and presentations had lower ROSES-S adherence compared to peer-reviewed journal articles. This may in part be attributable to limited reporting space in shorter form publication platforms like media reports and conference abstracts. Although the ROSES-S guideline was designed for full length seroepidemiologic study articles, grey literature including news media or public health reports can be used in evidence synthesis and, therefore, may warrant development of an adapted guideline for short form articles. Meanwhile, the lack of publication standards and peer-review for institutional reports may be contributing factors for poor reporting. The existing ROSES-S guideline is a suitable resource for those types of articles and could be better utilized.

Our results showed that risk of bias was associated with adherence to the ROSES-S guideline, similar to other studies that have found correlations between lower risk of bias and reporting completeness.^37, 38^ This association may arise from differences in the behaviors, traits, or circumstances of investigators that produce studies of higher versus lower methodological quality. For example, investigators conducting studies of higher methodological quality may have more resources and time to produce comprehensive study reports, or more training and knowledge about study methods, including reporting. If so, enhanced training for investigators conducting seroepidemiologic studies may be needed or additional resources to enable comprehensive reporting (e.g. user-friendly templates).

This study had two key strengths. Firstly, to the best of our knowledge, this was the first analysis to evaluate the reporting completeness of SARS-CoV-2 seroepidemiologic studies according to the ROSES-S guideline. Secondly, we sampled from the largest global repository of SARS-CoV-2 seroepidemiologic studies which was developed using a robust systematic review, thus the results likely have good generalizability.

This study had several limitations. Firstly, the 154 day lag time utilized in the analysis may not have been sufficiently long. Articles published after December 2021 may have been submitted before publication of the ROSES-S guideline and therefore subject to misclassification bias as the guideline was not available when the authors were reporting their data. However, our sensitivity analysis using a longer time delay (255 days, the third quartile of previously cited median publication lag time)^23^ did not differ from the main analysis (Appendix Table 3). Longer delays to publication are possible but likely only affected a small number of studies.^23^ Secondly, as some reporting criterion needed to be reported together to be interpretable, we may have overestimated adherence to multi-conditional items given that we scored the item as “reported” if any criterion were satisfied. Thirdly, decisions on whether a reporting item was achieved were subjective and other investigators may evaluate adherence differently. However, we mitigated this by developing our scoring sheet items in consultation with developers of the ROSES-S guideline, piloted our scoring, and discussed discrepancies to ensure inter-rater agreement.

In summary, this study found that the completeness of reporting in SARS-CoV-2 seroepidemiologic studies was suboptimal and associated with key study characteristics including article source of publication, risk of bias, and sampling method. Publication of the ROSES-S guideline was not associated with changes in reporting practices. Improvements in the completeness of reporting may serve to increase the transparency, comparability, and reproducibility of seroepidemiological research. Authors, journal editors, funders, decision-makers, and other stakeholders in infectious disease and public health should consider approaches to facilitate guideline adherence such as ROSES-S guideline endorsement and promotion, journal policies on reporting, and other potential funding incentives.

## Author contributions

**Brianna Cheng:** Conceptualization; investigation; methodology; project administration; supervision; writing – original draft (supporting); writing – review and editing (equal). **Emma Loeschnik:** Investigation; methodology; project administration; supervision; writing – original draft (supporting); writing – review and editing (equal). **Anabel Selemon:** Investigation; methodology; project administration; writing – original draft (supporting); writing – review and editing (equal). **Reza Hosseini:** Data curation; formal analysis; visualization; writing – original draft (supporting); writing – review and editing (equal). **Jane Yuan:** Data curation; formal analysis; visualization; writing – original draft (supporting); writing – review and editing (equal). **Harriet Ware**: Formal analysis; visualization; writing – original draft (supporting); writing – review and editing (equal). **Xiaomeng Ma:** Methodology; writing – original draft (supporting); writing – review and editing (equal). **Christian Cao:** Data curation; writing – original draft (supporting); writing – review and editing (equal). **Isabel Bergeri:** Conceptualization; methodology; writing – original draft (supporting); writing – review and editing (equal). **Lorenzo Subissi:** Methodology; writing – original draft (supporting); writing – review and editing (equal). **Hannah C Lewis:** Methodology; writing – original draft (supporting); writing – review and editing (equal). **Tyler Williamson:** Supervision; writing – review and editing (equal). **Paul Ronksley:** Supervision; writing – review and editing (equal). **Rahul K Arora:** Conceptualization; methodology; funding acquisition; project administration; supervision; writing – original draft (supporting); writing – review and editing (equal). **Mairead Whelan:** Project administration; funding acquisition; resources; supervision; writing – original draft (supporting); writing – review and editing (equal). **Niklas Bobrovitz:** Conceptualization; methodology; funding acquisition; project administration; supervision; writing – original draft (supporting); writing – review and editing (equal).

## Data Availability

Findings of this study are available within the article and its supplementary materials. The corresponding author may be contacted for all other requests.

## Funding statement

SeroTracker receives funding for SARS-CoV-2 seroprevalence study evidence synthesis from the Public Health Agency of Canada through Canada’s COVID-19 Immunity Task Force, the World Health Organization Health Emergencies Programme, the Robert Koch Institute, and the Canadian Medical Association Joule Innovation Fund.

## Conflict of interest statement

No funding source had any role in the design of this study, its execution, analyses, interpretation of the data, or decision to submit results. This manuscript does not necessarily reflect the views of the World Health Organization or any other funders.

## Ethics statement

This study did not require ethics approval.

## Notes

### Clinical Protocols

https://www.crd.york.ac.uk/prospero/display_record.php?ID=CRD42020183634

