## Appendix for "Adherence of SARS-CoV-2 seroepidemiologic studies to the ROSES-S reporting guideline during the COVID-19 pandemic"

**Figure 1: PRISMA flow diagram of inclusion**

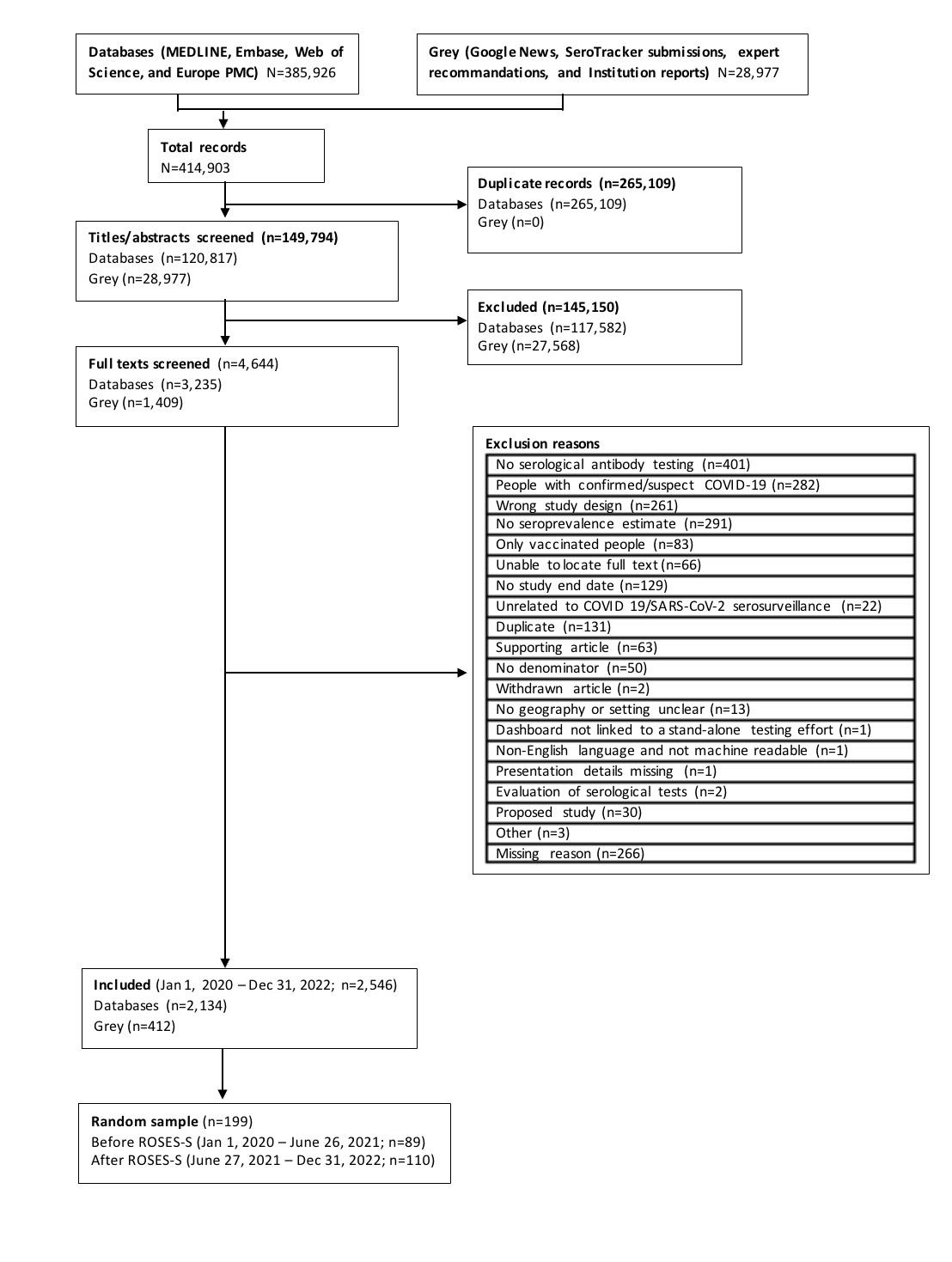

**Figure 2: Adherence to ROSES-S (%) overall and per domain**

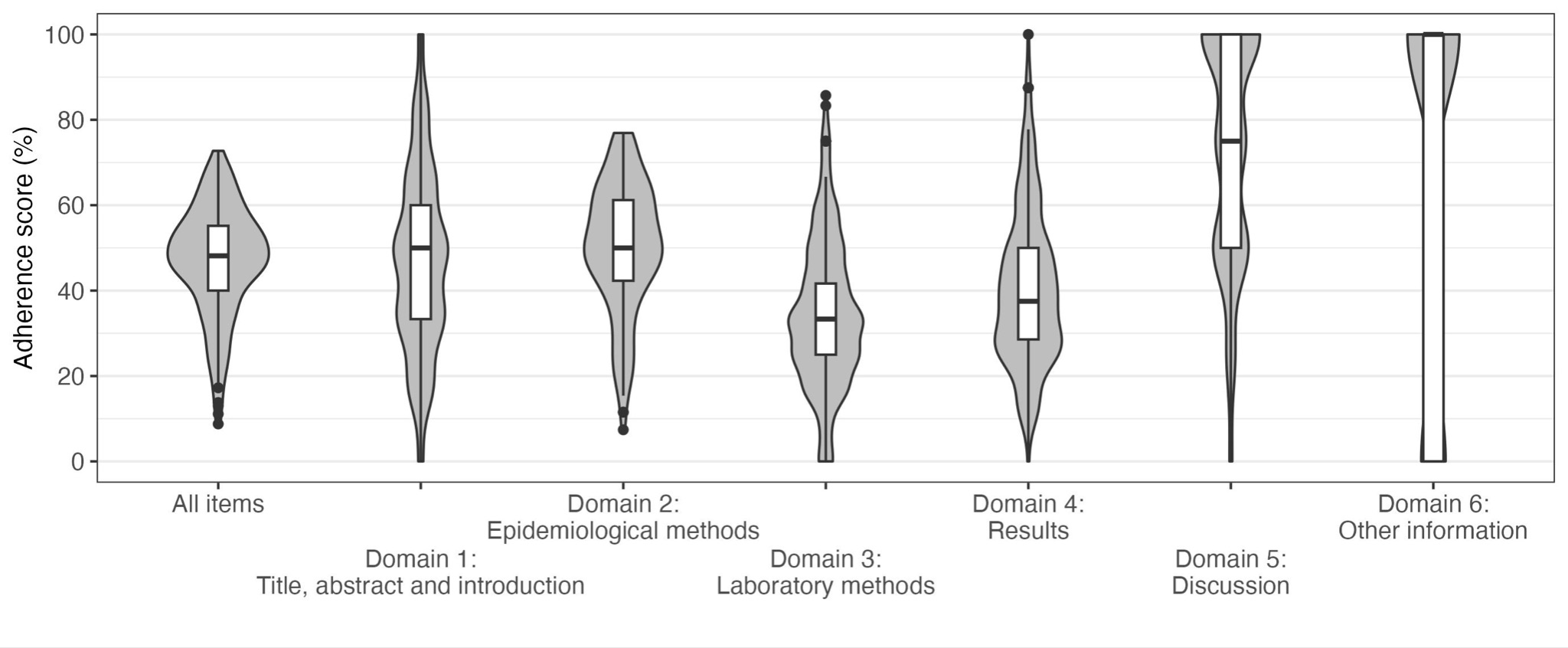

**Legend**

⇾ For all violin plots, n=199, except for Domain 6, for which n=197.

**Table 1: Risk of bias of included studies based on the modified version of the Joanna Briggs Institute (JBI) checklist**

| **Variable** | **Description** | **Overall** | **ROSES-S publication** | | |
| --- | --- | --- | --- | --- | --- |
|  |  |  | **Before** | **0-154 days after** | **155-553 days after** |
| **N** |  | 199 | 89 | 50 | 60 |
| **Overall Risk of Bias n/N (%)** | | | | | |
| High |  | 123/199 (61.8) | 61/89 (68.5) | 26/50 (52.0) | 36/60 (60.0) |
| Moderate |  | 66/199 (33.2) | 25/89 (28.1) | 22/50 (44.0) | 19/60 (31.7) |
| Low |  | 10/199 (5.0) | 3/89 (3.4) | 2/50 (4.0) | 5/60 (8.3) |
| **Modified JBI Item = Yes n/N (%)** | | | | | |
| JBI 1 | Was the sample frame appropriate to address the target population? | 85/199 (42.7) | 36/89 (40.4) | 21/50 (42.0) | 28/60 (46.7) |
| JBI 2 | Were study participants recruited in an appropriate way? | 40/199 (20.1) | 14/89 (15.7) | 10/50 (20.0) | 16/60 (26.7) |
| JBI 3 | Was the sample size adequate? | 125/199 (62.8) | 57/89 (64.0) | 35/50 (70.0) | 33/60 (55.0) |
| JBI 4 | Were the study subjects and the setting described in detail? | 121/199 (60.8) | 47/89 (52.8) | 41/50 (82.0) | 33/60 (55.0) |
| JBI 5 | Was data analysis conducted with sufficient coverage of the identified sample? | 60/199 (30.2) | 29/89 (32.6) | 12/50 (24.0) | 19/60 (31.7) |
| JBI 6 | Were valid methods used for the identification of the condition? | 102/199 (51.3) | 44/89 (49.4) | 25/50 (50.0) | 33/60 (55.0) |
| JBI 7 | Was the condition measured in a standard, reliable way for all participants? | 199/199 (100.0) | 89/89 (100.0) | 50/50 (100.0) | 60/60 (100.0) |
| JBI 8a | Was there appropriate adjustment for test characteristics? | 110/199 (55.3) | 50/89 (56.2) | 27/50 (54.0) | 33/60 (55.0) |
| JBI 8b | Was there appropriate adjustment for population characteristics? | 25/199 (12.6) | 9/89 (10.1) | 6/50 (12.0) | 10/60 (16.7) |
| JBI 9 | Was the response rate adequate, and if not, was the low response rate unlikely to introduce bias? | 37/199 (18.6) | 18/89 (20.2) | 15/50 (30.0) | 4/60 (6.7) |
| Colour key, based on percentage of item being ‘Yes’: ■ 0.0-19.9; ■ 20.0-39.9; ■ 40.0-59.9; ■ 60.0-79.9; ■ 80.0-100.0. | | | | | |

### **Table 2: Median and IQR adherence scores of study characteristics stratified by ROSES-S publication time period**

| **Variable** | |  | **Overall** |  | **ROSES-S publication** | | | | |
| --- | --- | --- | --- | --- | --- | --- | --- | --- | --- |
|  |  |  |  |  | **Before** | **0-154 days after** | | **155-553 days after** | |
|  | | **n (%)** | **median [IQR]** | **n (%)** | **median [IQR]** | **n (%)** | **median [IQR]** | **n (%)** | **median [IQR]** |
| **Source Type** | |  |  |  |  |  |  |  |  |
| Journal article (Peer-Reviewed) | | 136 (68.3) | 49.1  [43.0, 54.8] | 57 (28.6) | 48.1  [40.4, 55.6] | 41 (20.6) | 50.9  [43.4, 54.4] | 38 (19.1) | 47.4  [43.9, 53.2] |
| Pre-print | | 39 (19.6) | 52.6  [45.9, 61.5] | 18 (9.0) | 51.8  [46.6, 60.5] | 7 (3.5) | 55.2  [47.4, 56.7] | 14 (7.0) | 53  [45.6, 62.7] |
| Institutional report | | 17 (8.5) | 38.6  [28.8, 45.3] | 12 (6.0) | 33.8  [24.5, 40.4] | 2 (1.0) | 48.1  [47.2, 49.1] | 3 (1.5) | 38.6  [35.4, 46.5] |
| Conference, Abstract or News | | 7 (3.5) | 22  [15.1, 23.7] | 2 (1.0) | 12  [11.6, 12.5] | - | - | 5 (2.5) | 22.8  [22.0, 24.6] |
| **Study Design** | |  |  |  |  |  |  |  |  |
| Cross-sectional survey | | 149 (74.9) | 47.3  [38.0, 55.6] | 72 (36.2) | 46.8  [34.5, 55.8] | 37 (18.6) | 51.7  [45.5, 56.4] | 40 (20.1) | 45.6  [36.2, 52.8] |
| Repeated cross-sectional survey | | 21 (10.6) | 47.4  [45.6, 53.4] | 7 (3.5) | 46.4  [39.3, 50.4] | 2 (1.0) | 48.1  [47.2, 49.1] | 12 (6.0) | 49.6  [46.2, 54.4] |
| Cohort study | | 29 (14.6) | 50.9  [43.9, 56.1] | 10 (5.0) | 52.8  [46.2, 60.4] | 11 (5.5) | 50.8  [41.0, 53.1] | 8 (4.0) | 51.3  [43.9, 57.8] |
| **Sampling Frame** | |  |  |  |  |  |  |  |  |
| General population^†^ | | 105 (52.8) | 47.1  [38.0, 54.4] | 43 (21.6) | 44.4  [27.8, 52.2] | 18 (9.0) | 57.8  [47.2, 56.2] | 44 (22.1) | 47.4  [39.1, 54.6] |
| Special population | | 94 (47.2) | 49.1  [43.9, 56.3] | 46 (23.1) | 50.9  [44.0, 59.6] | 32 (16.1) | 49.5  [43.3, 54.6] | 16 (8.0) | 46.6  [43.9, 54.8] |
| **Sampling Method** | |  |  |  |  |  |  |  |  |
| Non-Probability | | 142 (71.3) | 48.1  [42.7, 55.1] | 66 (33.2) | 47.7  [42.7, 56.6] | 36 (18.1) | 50.4  [44.9, 54.6] | 40 (20.1) | 46.1  [39.4, 52.8] |
| Probability | | 40 (20.1) | 51.3  [45.6, 59.6] | 14 (7.0) | 49.1  [38.6, 55.3] | 10 (5.0) | 52.3  [50.2, 59.2] | 16 (8.0) | 50.9  [45.8, 61.7] |
| Unclear | | 17 (8.5) | 34.5  [22.0, 50.0] | 9 (4.5) | 23.2  [17.6, 34.6] | 4 (2.0) | 46.7  [41.2, 51.6] | 4 (2.0) | 31.8  [26.9, 40.4] |
| **Sample Size** | |  |  |  |  |  |  |  |  |
| < 500 | | 63 (31.7) | 46.6  [39.5, 52.8] | 28 (14.1) | 47.7  [39.9, 52.7] | 17 (8.5) | 48.1  [41.4, 52.6] | 18 (9.0) | 44.7  [37.9, 55.0] |
| 500-999 | | 44 (22.1) | 46.5  [35.0, 56.7] | 20 (10.1) | 45.9  [32.7, 59.8] | 10 (5.0) | 51.4  [40.1, 55.0] | 14 (7.0) | 46.5  [35.4, 54.8] |
| 1000+ | | 92 (46.2) | 50  [43.6, 56.4] | 41 (20.6) | 48.1  [38.9, 56.6] | 23 (11.6) | 51.7  [48.6, 57.1] | 28 (14.1) | 49.1  [43.6, 53.7] |
| **Overall JBI** | |  |  |  |  |  |  |  |  |
| High | | 123 (61.8) | 46.6  [36.9, 52.8] | 61 (30.7) | 46.6  [36.5, 52.8] | 26 (13.1) | 48.6  [42.1, 52.5] | 36 (18.1) | 45.6  [35.9, 51.3] |
| Moderate | | 66 (33.2) | 52.7  [45.3, 59.4] | 25 (12.6) | 52.8  [42.9, 59.6] | 22 (11.1) | 54.8  [49.3, 59.4] | 19 (9.5) | 50.9  [43.4, 54.8] |
| Low | | 10 (5.0) | 48.1  [38.6, 58.6] | 3 (1.5) | 38.5  [32.6, 38.7] | 2 (1.0) | 48.1  [47.2, 49.1] | 5 (2.5) | 61.4  [50.0, 62.5] |
| **Geographic Scope** | |  |  |  |  |  |  |  |  |
| National | | 42 (21.1) | 49.1  [39.1, 52.8] | 17 (8.5) | 39.6  [36.8, 52.6] | 11 (5.5) | 50.9  [48.1, 54.0] | 14 (7.0) | 50.4  [46.1, 53.2] |
| Regional | | 35 (17.6) | 51.9  [43.9, 61.3] | 12 (6.0) | 58.5  [51.4, 65.6] | 8 (4.0) | 53.2  [50.4, 60.2] | 15 (7.5) | 43.9  [33.0, 51.8] |
| Local | | 122 (61.3) | 47.3  [40.6, 55.1] | 60 (30.2) | 47.2  [37.1, 53.3] | 31 (15.6) | 49.1  [42.5, 54.8] | 31 (15.6) | 46.6  [41.4, 55.2] |
| **HRP Status** | |  |  |  |  |  |  |  |  |
| HRP | | 37 (18.6) | 51.9  [42.9, 56.1] | 14 (7.0) | 44  [41.0, 54.2] | 5 (2.5) | 56.4  [52.8, 59.6] | 18 (9.0) | 51.3  [43.1, 55.9] |
| Non-HRP | | 162 (81.4) | 47.8  [39.6, 55.0] | 75 (37.7) | 44  [41.0, 54.2] | 45 (22.6) | 56.4  [52.8, 59.6] | 42 (21.1) | 51.3  [43.1, 55.9] |
| **WHO Unity Protocol^‡^ ,** | |  |  |  |  |  |  |  |  |
| Unity-Aligned | | 44 (22.1) | 49.1  [42.8, 53.7] | 18 (9.0) | 44.9  [38.6, 47.9] | 12 (6.0) | 51.9  [49.1, 59.7] | 14 (7.0) | 50.9  [46.4, 54.4] |
| Not Unity-Aligned | | 155 (77.9) | 48.1  [39.5, 55.6] | 71 (35.7 | 49.1  [37.9, 57.5] | 38 (19.1) | 50.4  [43.9, 55.0] | 46 (23.1) | 45.6  [36.8, 55.0] |
| **Citation of Reporting Guideline** | |  |  |  |  |  |  |  |  |
| Yes |  | 6 (3.0) | 57  [48.9, 59.2] | 2 (1.0) | 63.5  [61.6, 65.4] | 1 (0.5) | 57.9  [57.9, 57.9] | 3 (1.5) | 46.6  [46.1, 51.3] |
|  | STROBE | 5 (2.5) | 57.9  [56.1, 59.6] | 2 (1.0) | 63.5  [61.6, 65.4] | 1 (0.5) | 57.9  [57.9, 57.9] | 2 (1.0) | 51.3  [48.9, 53.7] |
|  | TREND | 1 (0.5) | 45.6  [45.6, 45.6] | - | - | - | - | 1 (0.5) | 45.6  [45.6, 45.6] |
| No |  | 193 (97.0) | 48.1  [39.6, 55.2] | 87 (43.7) | 47.1  [37.9, 55.1] | 49 (24.6) | 50.8  [45.5, 55.2] | 57 (28.6) | 46.6  [39.3, 54.4] |

† General population includes the following sampling frames: household and community samples, blood donors, residual sera, persons living in slums, pregnant or parturient women, and representative patient populations.

‡ WHO Unity aligned: studies aligned with the WHO Unity protocol.

Abbreviations: HRP, Humanitarian Response Plan; STROBE, Strengthening the Reporting of Observational Studies in Epidemiology; TREND, Transparent Reporting of Evaluations with Nonrandomized Designs.

**Table 3:** [**Univariable beta regression separately evaluating the association of each candidate predictor with total adherence score**](https://docs.google.com/document/d/11yXuNCT-TiidV5NIvTnMY_vtMzfUXoyg/edit#heading=h.2s8eyo1)

| **Variable** |  | **Model coefficients** | | | **Marginal effects (%)** | | |
| --- | --- | --- | --- | --- | --- | --- | --- |
|  |  | **Estimate (95% CI)** | **p** |  | **Estimate (95% CI)** | **p** |  |
| Publication date, compared to ROSES-S (month); 154 day lag^†^ | (Intercept) | 49.58 (-409.43 - 508.59) |  |  |  |  |  |
|  | Before | 0.00 (-0.03 - 0.02) | 0.815 |  | -0.07 ( -0.69 - 0.54) | 0.815 |  |
|  | 0-154 days after | 0.04 (-0.03 - 0.12) | 0.256 |  | 1.11 ( -0.82 - 3.03) | 0.256 |  |
|  | 155-553 days after | -0.06 (-0.14 - 0.02) | 0.126 |  | -1.53 ( -3.51 - 0.44) | 0.126 |  |
| Publication date, compared to ROSES-S (month); 255 day lag^†^ | (Intercept) | -17.12 (-443.12 - 408.89) |  |  |  |  |  |
|  | Before | 0.00 (-0.02 - 0.02) | 0.955 |  | 0.02 ( -0.56 - 0.59) | 0.955 |  |
|  | 0-255 days after | 0.02 (-0.03 - 0.07) | 0.463 |  | 0.50 ( -0.84 - 1.83) | 0.463 |  |
|  | 256-553 days after | -0.05 (-0.12 - 0.02) | 0.166 |  | -1.26 ( -3.06 - 0.54) | 0.166 |  |
| WHO HRP status | (Intercept) | -0.15 (-0.23 - -0.07) |  |  |  |  |  |
|  | Non-HRP | Reference |  |  | Reference |  |  |
|  | HRP | 0.09 (-0.10 - 0.28) | 0.351 |  | 2.23 ( -2.49 - 6.96) | 0.352 |  |
| WHO Unity alignment | (Intercept) | -0.14 (-0.22 - -0.05) |  |  |  |  |  |
|  | Not Unity-aligned | Reference |  |  | Reference |  |  |
|  | Unity-aligned | 0.03 (-0.14 - 0.21) | 0.719 |  | 0.81 ( -3.63 - 5.24) | 0.720 |  |
| Sampling frame | (Intercept) | -0.21 (-0.31 - -0.11) |  |  |  |  |  |
|  | General population^‡^ | Reference |  |  | Reference |  |  |
|  | Special population | 0.17 (0.02 - 0.31) | 0.024 | * | 4.16 ( 0.52 - 7.80) | 0.024 | * |
| Geographic scope | (Intercept) | -0.16 (-0.25 - -0.06) |  |  |  |  |  |
|  | Local | Reference |  |  | Reference |  |  |
|  | Regional | 0.13 (-0.07 - 0.33) | 0.191 |  | 3.29 ( -1.68 - 8.25) | 0.192 |  |
|  | National | 0.01 (-0.17 - 0.20) | 0.878 |  | 0.36 ( -4.27 - 4.99) | 0.878 |  |
| Study design | (Intercept) | -0.15 (-0.24 - -0.07) |  |  |  |  |  |
|  | Cross-sectional survey | Reference |  |  | Reference |  |  |
|  | Repeated cross-sectional study | 0.06 (-0.18 - 0.30) | 0.607 |  | 1.57 ( -4.47 - 7.61) | 0.607 |  |
|  | Cohort study | 0.12 (-0.09 - 0.33) | 0.249 |  | 3.07 ( -2.19 - 8.33) | 0.249 |  |
| Sample size | (Intercept) | -0.13 (-0.21 - -0.06) |  |  |  |  |  |
|  | Sample size | 0.00 (0.00 - 0.00) | 0.777 |  | 0.00 ( 0.00 - 0.00) | 0.777 |  |
| Overall risk of bias | (Intercept) | -0.23 (-0.32 - -0.14) |  |  |  |  |  |
|  | High | Reference |  |  | Reference |  |  |
|  | Moderate | 0.27 (0.12 - 0.43) | <0.001 | *** | 6.82 ( 2.96 - 10.67) | <0.001 | *** |
|  | Low | 0.10 (-0.23 - 0.43) | 0.544 |  | 2.56 ( -5.77 - 10.90) | 0.544 |  |
| Sampling method | (Intercept) | -0.10 (-0.18 - -0.02) |  |  |  |  |  |
|  | Non-probability | Reference |  |  | Reference |  |  |
|  | Probability | 0.10 (-0.08 - 0.27) | 0.277 |  | 2.43 ( -1.99 - 6.85) | 0.278 |  |
|  | Unclear | -0.58 (-0.85 - -0.32) | <0.001 | *** | -14.02 (-20.02 - -8.02) | <0.001 | *** |
| Source type | (Intercept) | -0.08 (-0.15 - 0.00) |  |  |  |  |  |
|  | Peer-reviewed journal article | Reference |  |  | Reference |  |  |
|  | Preprint | 0.20 (0.04 - 0.36) | 0.012 | * | 5.08 ( 1.07 - 9.08) | 0.012 | * |
|  | Institutional report | -0.55 (-0.79 - -0.32) | <0.001 | *** | -13.26 (-18.64 - -7.88) | <0.001 | *** |
|  | Conference abstract or news | -1.28 (-1.69 - -0.88) | <0.001 | *** | -27.59 (-34.30 - -20.87) | <0.001 | *** |
| Citation of any guidelines^§^ | (Intercept) | -0.14 (-0.21 - -0.07) |  |  |  |  |  |
|  | No | Reference |  |  | Reference |  |  |
|  | Yes | 0.35 (-0.08 - 0.78) | 0.108 |  | 8.76 ( -1.90 - 19.41) | 0.105 |  |
| ^†^ Publication date is a continuous variable with monthly increments. Each month is considered equal to 30.4 days. | | | | | | | |
| ^‡^ General population includes the following sampling frames: household and community samples, blood donors, residual sera, persons living in slums, pregnant or parturient women, and representative patient populations. | | | | | | | |
| ^§^ None cited ROSES-S. The only cited guidelines were STROBE (n=5) and TREND (n=1). | | | | | | | |
| Abbreviations: HRP, Humanitarian Response Plan. | | | | | | | |
