## Supplemental for "Adherence of SARS-CoV-2 seroepidemiologic studies to the ROSES-S reporting guideline during the COVID-19 pandemic"

**Supplementary file 1: ROSES-S scoring sheet**

| **Domain** | **Item** | | **Description** |
| --- | --- | --- | --- |
|  | **Title** | **#** |  |
| Domain 1: Title, abstract and introduction | Title and abstract | 1.1 | The term “seroepidemiologic,” “seroepidemiology,” “seroprevalence,” or “seroincidence” should be applied to the study in the title and abstract, and the medical subject heading “Seroepidemiologic Studies” be used when the report is of a population-based serological survey. |
|  |  | 1.2 | Provide a structured summary including, as applicable: objectives; population level (ie, national, regional, local), study design, study period, eligibility criteria of study participants, sampling dates and method, sample size, laboratory methods (assay used), results: seroprevalence and 95% CI, study limitations, conclusions and implications of key findings |
|  | Introduction | 2.1 | State what is known about the kinetics of antibody rise, decay, and persistence following SARS-CoV-2 infection, in the particular study setting/population, if possible. |
|  |  | 2.2 | State which SARS-CoV-2 viruses are circulating, including any variants. |
|  |  | 2.3 | State what is known about the sensitivity and specificity of the antibody detection assay being used. |
|  |  | 3.1 | State specific objectives, including any prespecified hypotheses |
| Domain 2: Epidemiological methods | Study design | 4.1 | State which specific seroepidemiologic study design was chosen and why. |
|  | Setting | 5.1 | Describe the setting, locations, and sampling frame, including periods of recruitment, exposure, follow-up, and data collection. |
|  |  | 5.2 | Describe the timing of the biological sampling in relation to the disease epidemiology in the study population (the beginning, peak, and end of virus transmission). |
|  |  | 5.3 | Describe any vaccination efforts that have been undertaken. |
|  |  | 5.4 | Where known, describe the timing of biological sampling in individuals in relation to disease onset and to exposures of interest. |
|  |  | 5.5 | State the interval between sequential biological samples (serial cross-sectional or longitudinal studies), or specify whether only a single sample was collected (cross-sectional study). |
|  | Participants | 6.1 | For case‐ascertained transmission studies, describe the method of case ascertainment and criteria for defining a “case.” Describe methods of follow‐up. |
|  |  | 6.2 | For household‐ or institution‐based transmission studies, describe the definition of a household or the institution. Describe methods of follow‐up. |
|  |  | 6.3 | For outbreak investigations involving serologic sampling, describe the setting in which the cases were identified, for example, village/residential setting, occupational workplace. Describe methods of follow‐up. |
|  |  | 6.4 | For a cohort study, give the eligibility criteria, and the sources and methods of sampling of participants. Describe methods of follow‐up. |
|  |  | 6.5 | For a case‐control study, give the eligibility criteria, and the sources and methods of case ascertainment and control selection. Give the rationale for the choice of cases and controls. For matched studies, give matching criteria and the number of controls per case. |
|  |  | 6.6 | For a cross‐sectional study, give the eligibility criteria, and the sources and methods of selection of participants. |
|  | Variables | 7.1 | Clearly define all outcomes, exposures, predictors, potential confounders, and effect modifiers. |
|  |  | 7.2 | The median age and range for each exposure group should be reported. |
|  |  | 7.3 | Describe the vaccination status of participants (specify vaccination status, vaccine manufacturer, number of doses, and timing of vaccination in relationship to collection of serum), if applicable, to affect the outcome measures. If relevant, describe measures taken to identify and record immunization history. |
|  |  | 7.4 | Describe any known or potential immunological cross‐reactivity that may bias the outcome measures. |
|  |  | 7.5 | Describe illness definitions and methods for ascertaining the presence or absence of clinical illness in subjects. |
|  | Data sources/ measurement biases | 8.1 | For each variable of interest, give sources of data and details of methods of assessment (measurement). Describe comparability of assessment methods if there is more than one group. |
|  |  | 8.2 | Give information separately for cases and controls in case‐control studies and, if applicable, for exposed and unexposed groups in cohort and cross‐sectional studies). |
|  | Bias | 9.1 | Describe any efforts to address potential sources of bias. |
|  | Study size | 10.1 | Describe the baseline estimated seroprevalence or incidence of infection and cite published literature to support these estimates. |
|  |  | 10.2 | Explain the steps that led to the final sample size. Report the numbers of individuals at each stage of the study—the numbers potentially eligible, examined for eligibility, confirmed eligible, included in the study, completing follow‐up, and analyzed. |
|  | Quantitative variables | 11.1 | Explain how quantitative variables were handled in the analyses. If applicable, describe which groupings were chosen and why. |
|  |  | 11.2 | Describe the serological assay's limit of detection and how this limit is defined or calculated. Describe how samples with a result below or on the borderline of the limit were handled in the analysis. |
|  |  | 11.3 | Define “seropositivity,” or the antibody titer change or change in other assay result used to define “seroconversion.” Avoid the term “seroconversion” unless referring to change from undetectable to detectable antibody level. Avoid the term “infection” but report “seroprevalence at a titer of ….”. |
|  | Statistical methods | 12.1 | Describe all statistical methods, including those used to control for confounding. |
|  |  | 12.2 | Describe any methods used to examine subgroups and interactions. |
|  |  | 12.3 | Describe all methods used to address sampling and selection biases (eg, weighting results, multilevel regression and post‐stratification). |
|  |  | 12.4 | Explain how missing data were addressed. |
|  |  | 12.5 | For a cohort study, explain how loss to follow‐up was addressed, if applicable. |
|  |  | 12.6 | For a case‐control study, explain how variables on which cases and controls were matched, if applicable. |
|  |  | 12.7 | For a cross‐sectional study, describe analytical methods taking account of sampling strategy, if applicable. |
|  |  | 12.8 | Describe any sensitivity analyses. |
|  |  | 12.9 | If relevant, report methods used to account for adjustment for assay performance (sensitivity and specificity), the probability of seropositivity or seroconversion if infected, and to account for decay in antibody titers over time. |
| Domain 3: Laboratory methods | Sample type and handling | 13.1 | Describe the sample type—whole blood, dried blood, serum or plasma. If plasma is used, specify the anticoagulant used (heparin, sodium citrate, EDTA, etc). |
|  |  | 13.2 | Describe the specimen storage conditions (4°C, −20°C, −80°C). If frozen prior to the analysis, describe the time to freezing and the number of freeze/thaw cycles prior to testing. |
|  | Serological assays | 13.3 | Wherever possible, use defined and standardized methods that have been established in more than one laboratory, and that ideally are commercially available in more than one country. Avoid laboratory‐level formulations if standardized formulations are available for the same analytical targets. |
|  |  | 13.4 | Specify the testing algorithm (if more than one test used) and assay type (eg, virus neutralization/microneutralization/surrogate neutralization; ELISA; LFIA; CLIA; other) and readout used to determine the endpoint titer. |
|  |  | 13.5 | Reference a previously published protocol, if used, and any modifications of the protocol. If a previously published protocol was not used, provide full details in supplementary materials. For in‐house assays, include a description of the assay format (e.g., direct or indirect immunoassay) as well as description of cutoff determination and which antibody isotype is targeted, and reference previously published validation data. |
|  |  | 13.6 | State what is known about the determinants of the variability of the antibody detection assay being used. |
|  |  | 13.7 | Specify the antigen(s) and antibody isotope target used, with standardized nomenclature and reference; specify whether live virus or pseudo virus was used (where applicable). Describe how the cutoff was established. If viral antigen produced in‐house is used, specify sequence, expression system (bacteria or mammalian cells). Specify reactivity with other coronavirus antigens (MERS‐CoV, SARS‐CoV, seasonal CoVs) in the same population. |
|  |  | 13.8 | Describe positive and negative controls used. Specify international standards used, if appropriate. |
|  |  | 13.9 | Describe starting and end dilutions. |
|  |  | 13.10 | Specify laboratory biosafety conditions. |
|  |  | 13.11 | Specify whether replication was performed, and if so, the acceptable replication parameters. |
|  |  | 13.12 | Specify whether a confirmatory assay was performed and all specifics of this assay, at the same level of detail. |
| Domain 4: Results | Participants | 13.13 | Report the numbers of individuals at each stage of the study—the numbers potentially eligible, examined for eligibility, confirmed eligible, included in the study, completing follow‐up, and analyzed. |
|  |  | 13.14 | Give reasons for non‐participation at each stage. Consider use of a flow diagram |
|  | Descriptive data | 14.1 | Give characteristics of study participants (eg, demographic, clinical, social) and information on exposures and potential risk factors for all participants, not solely stratified by outcome status. |
|  |  | 14.2 | Indicate the number of participants with missing data for each variable of interest. |
|  |  | 14.3 | For a cohort study, detail follow‐up time (eg, average and total amount). |
|  | Outcome data | 15.1 | For a cohort study, report the numbers of outcome events or summary measures over time. |
|  |  | 15.2 | For a case‐control study, report the numbers in each exposure category, or summary measures of exposure. |
|  |  | 15.3 | For a cross‐sectional study, report the numbers of outcome events or summary measures. |
|  | Main result | 16.1 | Report unadjusted estimates of distribution of seropositivity by age group. |
|  |  | 16.2 | Report methods to standardize the results from the study sample to the target population. |
|  | Other analyses | 17.1 | Report other analyses performed—analyses of subgroups and interactions, and sensitivity analyses. |
| Domain 5: Discussion | Key results | 18.1 | Summarize key results with reference to study objectives. |
|  | Limitations | 19.1 | Discuss limitations and strengths of the study. |
|  | Interpretation | 20.1 | Discuss the interpretation of the results in the context of known or potential cross‐reactivity, assay performance and other sources of bias |
|  | Generalizability | 21.1 | Discuss the generalizability (external validity) of the study results. |
| Domain 6: Other information | Ethics approval | 22.1 | Specify if institutional review board approval was received; if not, specify reason (eg, public health outbreak response/non‐research designation). |

### **Supplementary file 2: Beta regression methods**

Beta regression was performed using the betareg package. Univariable models were constructed for all candidate predictors. Among them, publication date was used in a continuous piecewise model with two knots, one at the date of ROSES-S publication and the other 154 days later, to assess trends of adherence to ROSES-S over time. This model was compared to a non-segmented model using the likelihood-ratio test. The same analysis was repeated with a publication time lag of 255 days for its second knot as a sensitivity analysis. Publication date was entered in these models as a date variable and its coefficients were multiplied by 30.4 to represent monthly increments.

A two-step strategy was used to fit the mean and the precision beta regression sub-models. The mean component was fit first, assuming constant dispersion. The MuMIN package was used for a best subset model building approach with AIC as the selection criterion. When multiple models were within two delta AIC of the top model, the most parsimonious one was chosen. Then, the finalized mean sub-model was held constant while selecting the regressors for the precision sub-model using the same best subset approach. Finally, marginal effects with 95% confidence intervals were calculated using the mfx package to facilitate the interpretation of average change in total adherence scores at percentage level.

### **Supplementary file 3: Reference list of included studies**

49. Canadian Blood Services. *COVID-19 Seroprevalence Report October 13, 2021 Report #12 - July 2021 Survey Tracking Seroprevalence in the Vaccine Era*.; 2021.

133. Canadian Blood Services. *COVID-19 Seroprevalence Report October 13, 2021 Report #13 - August 2021 Survey Tracking Seroprevalence in the Vaccine Era*.; 2021.

139. Canadian Blood Services. *COVID-19 Seroprevalence Report April 10th, 2022 Report #19*.; 2022. [N/A](https://n/A).
